# Low cost and real-time surveillance of enteric infection and diarrhoeal disease using rapid diagnostic tests: A pilot study

**DOI:** 10.1101/2022.03.07.22271752

**Authors:** Samuel I. Watson, Mohammed Atique Ul Alam, Ryan T.T. Rego, Richard J. Lilford, Ashok Kumar Barman, Baharul Alam, A.S.G. Faruque, Md. Sirajul Islam

## Abstract

**Background:** Real-time disease surveillance is an important component of infection control in at-risk populations. However, data on cases or from lab testing is often not available in many low-resource settings. Rapid diagnostic tests (RDT), including immunochromatographic assays, may provide a low cost, expedited source of infection data.

**Methods:** We conducted a pilot survey-based prevalence mapping study of enteric infection in Camp 24 of the camps for the forcibly displaced Rohingya population from Myanmar in Cox’s Bazar, Bangladesh. We randomly sampled the population and collected and tested stool from under-fives for eight pathogens using RDTs in January-March 2021 and September-October 2021. A Bayesian geospatial statistical model allowing for imperfect sensitivity and specificity of the tests was adapted.

**Results:** We collected and tested 396 and 181 stools in the two data collection rounds. Corrected prevalence estimates ranged from 0.5% (Norovirus) to 27.4% (Giardia). Prevalence of *E*.*coli* O157, Campylobacter, and Cryptosporidium were predicted to be higher in the high density area of the camp with relatively high probability (70-95%), while Adenovirus, Norovirus, and Rotavirus were lower in the areas with high water chlorination. Clustering of cases of Giardia and Shigella was also observed, although associated with relatively high uncertainty.

**Conclusions:** With an appropriate correction for diagnostic performance RDTs can be used to generate reliable prevalence estimates, maps, and well-calibrated uncertainty estimates at a significantly lower cost than lab-based studies, providing a useful approach for disease surveillance in these settings.

## INTRODUCTION

Real-time surveillance is an important component of preventative interventions against infectious disease epidemics. The identification of emerging disease clusters can support the targeting of preventative measures to reduce disease transmission. Ratnayake et al, for example, review the use of case area-targeted intervention (CATI) for cholera.[1] In response to the early detection of a cluster, measures including chemoprophylaxis, water treatment and vaccination can be deployed to small areas.

The identification of disease clusters typically relies on the statistical modelling of georeferenced and time-stamped case data. The “Gold standard” case data is a large random sample of the population using PCR-based testing. However, PCR-based testing is expensive and requires laboratories with skilled technicians, which may be unavailable in many contexts. Alternative data sources with location and time information, such as hospital admissions or use of electronic health services, can be used to monitor disease spread in a population and model changes in incidence.[2, 3] Again though, this information may not be routinely available in many low resource settings. In the absence of geolocated case data or PCR-based surveys, surveys using rapid diagnostic tests (RDTs) may present a useful alternative for real-time disease surveillance.

RDTs are typically immunochromatographic assays that provide a visual response in the presence of specific antigens. These tests are relatively low cost, they can be administered by people with little training, and provide results within around 20 minutes (e.g.[4–7]) The performance of diagnostic tests is generally reported in terms of its sensitivity (the probability a person with the disease tests positive) and its specificity (the probability a person without the disease tests negative). With imperfect sensitivity and specificity, the crude test positive proportion is a biased estimator for the prevalence, and hence any derived measures like relative risk or odds ratios are also like to be biased.[8, 9] However, one can allow for the sensitivity and specificity in statistical analyses, which would produce unbiased estimators that appropriately reflect the additional uncertainty caused by the imperfect test.[8, 10]

When used and modelled appropriately RDTs may therefore be a useful tool for monitoring infection in a population. Indeed, their uses may extend to other applications including as an outcome measure in evaluations of interventions. For example, water, sanitation, and hygiene (WASH) interventions have been the subject of many large-scale trials in recent years.[11–14] Almost without exception though, these studies have used self-reported diarrhoea as their primary outcome. Diarrhoea is subject to many biases in its measurement and can be considered to have very poor “diagnostic performance” with respect to enteric infection, the transmission of which WASH interventions aim to prevent. PCR testing for enteric pathogens may be too costly or the infrastructure unavailable to include in such large trials. RDTs may therefore also provide a useful middle ground beyond surveillance applications.

In this study, we use RDTs to estimate and map the prevalence in the under-fives of several enteric pathogens at two time points in the camps for Forcibly Displaced Rohingya Population from Myanmar (FDRPM) in Cox’s Bazar, Bangladesh. These camps are typically densely populated and have inadequate WASH related facilities, leading to potentially high risk of diarrhoeal diseases. Our aim was to predict the prevalence of enteric infection of different pathogens and their spatial and temporal distribution in a FDRPM camp, and in so doing establish that RDTs could be used in such settings given the absence of previous research using them for this purpose, and develop the statistical methodology to incorporate uncertainty about the performance of the tests.

## METHODS

### Study setting

We conducted our study in Camp 24 of the FDRPM Camps in Cox’s Bazar, Bangladesh. Figure 1 shows a map of the camp. The camp consists of a densely populated area in the North-East with lower density settlements in the remaining area. WASH infrastructure has been slowly developed by several NGOs in the previous few years and generally consists of tube wells to provide water and improved latrines for sanitation.

**FIGURE 1.**
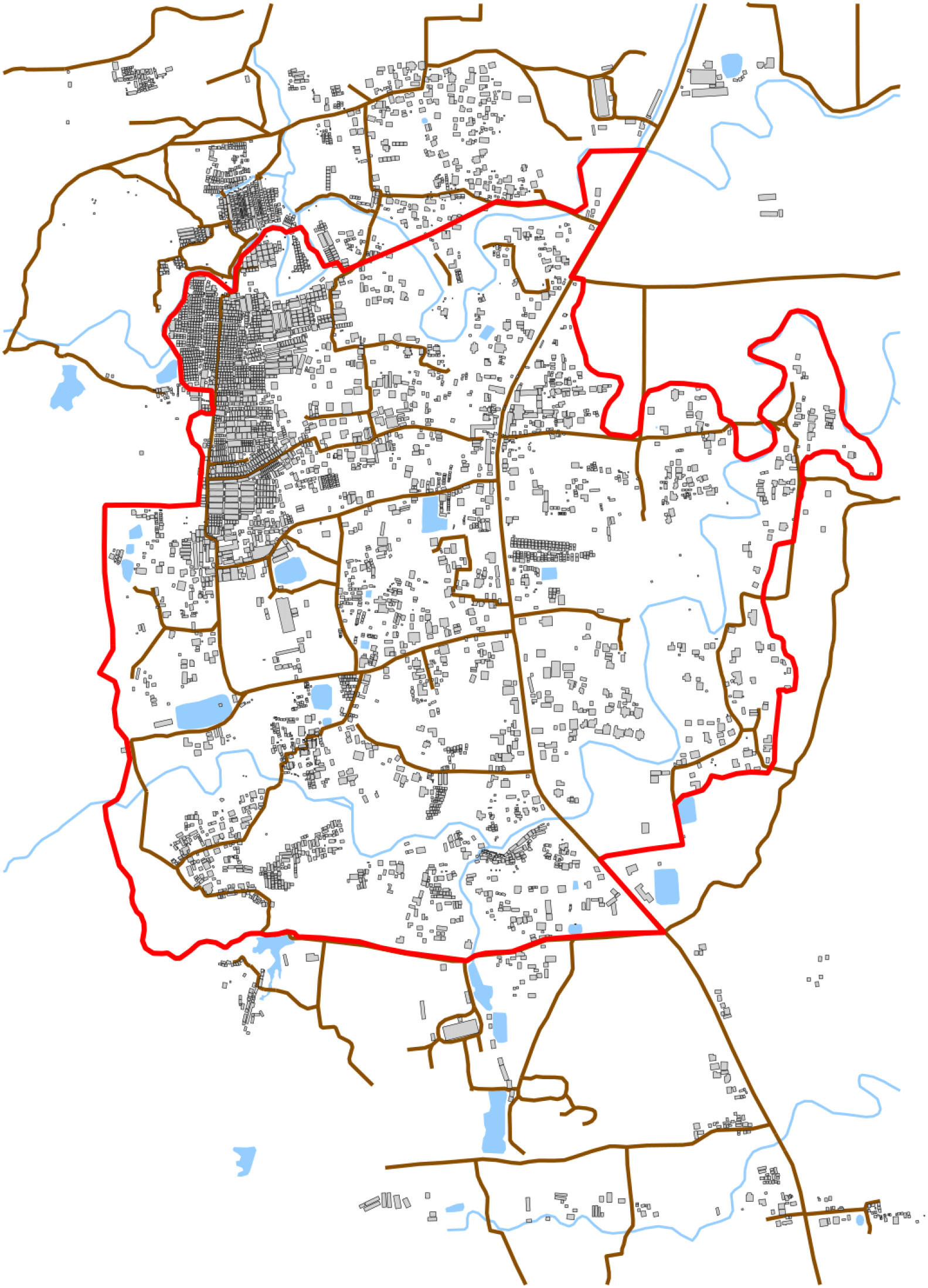
Map of Camp 24. The camp shows the structures in the camp reflecting the high density in the North-East of the camp. The boundary is indicated in red, the roads are brown, and water is blue.

### Sample

We aimed to include 400 households with a child between the age of 18 and 48 months in the study, with good dispersion across the area of the camp. We lacked a complete census for the camp that could identify eligible households. We therefore drew a sample from all households in the camp. Each sampled household was then visited and if they had a child under five years of age, we proceeded with the consent and interviewing process. The camp is divided into “blocks” and each residential location in each block is assigned a sequential “door number”, starting at one. We obtained the total number of households in each block and then sampled from the door numbers proportional to the block size. Based on previous work in the area we estimated that approximately one third of households would have a child under the age of five and the response rate would be close to 100%. We conducted two rounds of the survey in January-March 2021 (Round 1) and September-October 2021 (Round 2). We therefore sampled 1,200 households to obtain a sample of 400 in round 1. We aimed to revisit the same participating households in round 2.

### Survey and stool sampling

#### Household survey

At each participating household, we sought consent from the primary caregiver of the children under five. We then conducted a short survey capturing basic demographic and socioeconomic background data using the Open Data Kit software on tablet devices, including age, sex, level of education, time in Bangladesh, and water and sanitation facilities. We also captured the GPS location of the household in the camp.

#### Stool testing

A random child under five was sampled and the caregiver was provided with a plastic container with a barcode identifier for the child’s stool. The fieldworkers then returned the next day to collect the stool. A sample of the drinking water from the household was also collected from the container they normally used for testing the concentration of chlorine. The stool was taken to a field office in the camp. We used ProFlow tests produced by Pro-Lab Diagnostics, Wirral, United Kingdom for a set of eight pathogens listed in Table 1. Results from the tests were recorded in a survey form and linked to the household ID. The field worker also recorded whether the stool was diarrhoea or not.

**Table 1.** Summary of used rapid diagnostic tests and their reported sensitivity and specificity in the literature

| Pathogen | Reported sensitivity range | Prior | Reported specificity range | Prior | Refs |
| --- | --- | --- | --- | --- | --- |
| E. Coli O157 | 80 to 98% | Beta(18,2) | 95 to 100% | Beta(98,2) | [6] |
| Cryptosporidium | 50 to 100% | Beta(15,5) | 90 to 100% | Beta(95,5) | [7, 15] |
| Giardia | 50 to 95% | Beta(15,5) | 90 to 100% | Beta(95,5) | [7, 15] |
| Shigella | 80 to 95% | Beta(18,2) | 95 to 100% | Beta(98,2) | [16] |
| Campylobacter | 80 to 90% | Beta(17,3) | 95 to 100% | Beta(98,2) | [17, 18] |
| Rotavirus | 75 to 100% | Beta(17,3) | 98 to 100% | Beta(99,1) | [4, 5] |
| Norovirus | 90 to 100% | Beta(19,1) | 99 to 100% | Beta(99,1) | [5, 19] |
| Adenovirus | 90 to 100% | Beta(19,1) | 98 to 100% | Beta(99,1) | [20–22] |

### Data and analysis

#### Sensitivity and specificity

As stated, we planned to account for the uncertainty due to the performance of the diagnostic tests in our statistical analysis. Table 1 provides estimates of test performance in terms of the sensitivity and specificity for each test based on values reported in the literature. All of the tests had high reported sensitivity, however specificity was more variable.

#### Covariates

We included two spatially-referenced covariates in our statistical model. First, we used the density of structures on the map (Figure 1) as a proxy for population density. Second, we used the estimated level of water chlorination in parts per million (ppm). From each surveyed household’s water sample we tested the chlorine levels, these data were then smoothed over the area for each survey round using kernel density smoothing. Figure 2 shows the covariates within the boundary of the camp.

**Figure 2.**
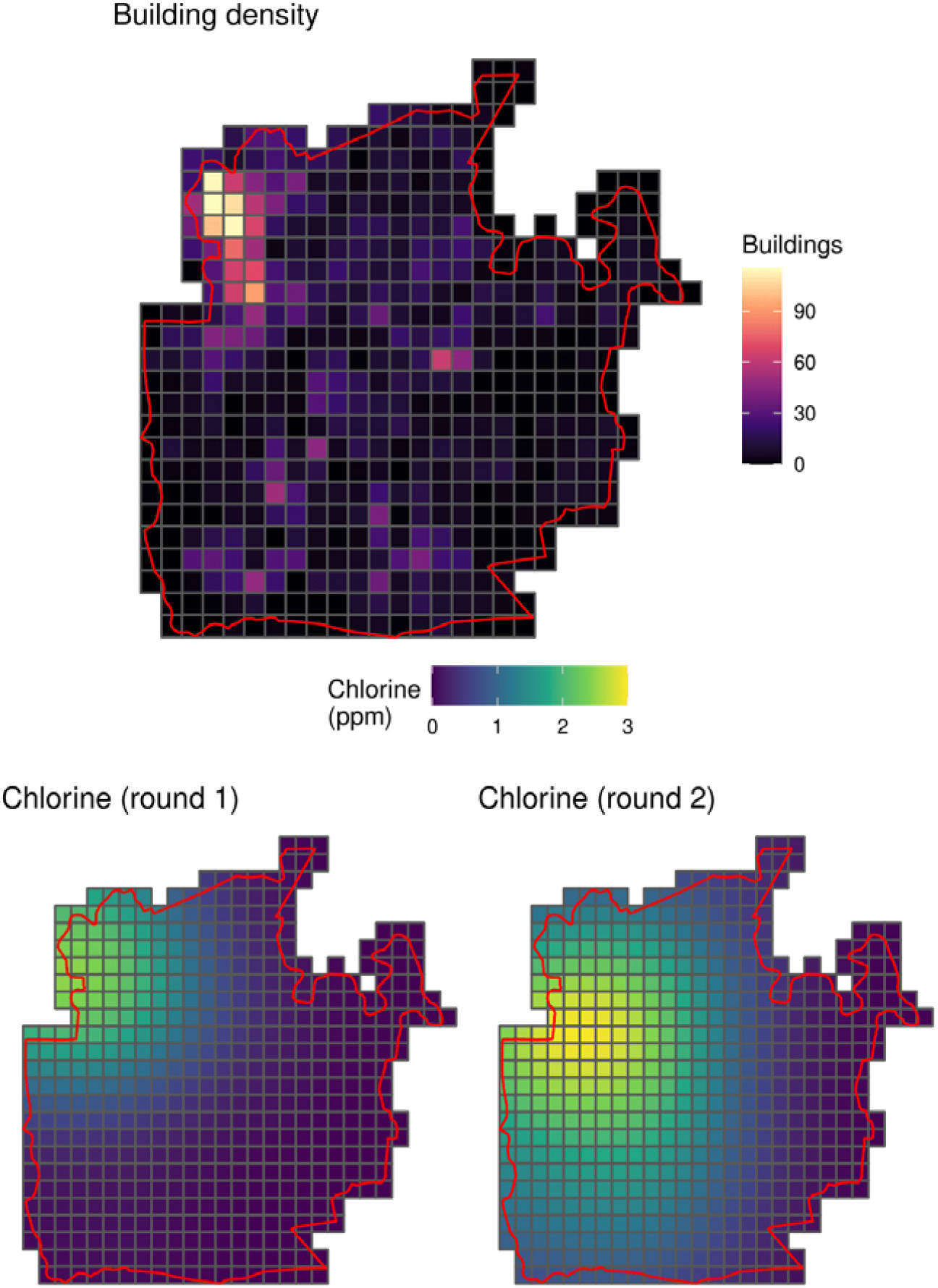
Spatially-referenced covariates in Camp 24: building density (top) and spatially-smoothed levels of chlorine in the drinking water supply in both survey rounds (bottom).

Inclusion of covariates can improve predictions and reduce uncertainty.[3, 23] We note that the parameters in geospatial statistical models may be biased and difficult to interpret,[24, 25] and so we do not aim to provide inference on the “effects” of either of the included covariates beyond their relative comparisons of their magnitudes in predicting the outcome.

#### Statistical model

A technical description of the methods is provided in the Supplementary Information. In brief, we specified a binomial geospatial statistical model. For a location *s* in our area of interest at time *t* = 1,2 we observe the outcome of the test for person *i* = 1, …, *N* as *y*(*s*_*i*_, *t*) where:

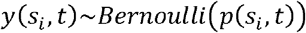

For each location and time we define the linear predictor:

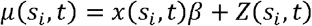

where *x*(*s, t*) are the spatially and temporally referenced covariates (Figure 2) and *Z*(*s, t*) is a smooth latent process over the area of interest, which we describe below. If we ignore the diagnostic performance of the tests then the model would have *p*(*s*_*i*_, *t*) = *h*^−1^ (*µ*(*s*_*i*_, *t*)) where *h*^−1^(.) is the inverse-logit function. We refer to this as the “uncorrected model”.

To take into account the sensitivity and specificity, the probability in the model should reflect the probability of testing positive, rather than the probability of having the disease. The test positive probability is:

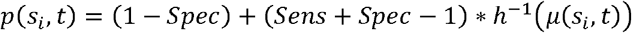

where *Sens* is the sensitivity and *Spec* is the specificity. We refer to this as the “corrected model”.

##### Prior distributions

The standard geospatial statistical model formulation specifies a Gaussian process prior for the term *Z*(*s, t*). We use an accurate approximation to a Gaussian process prior to improve computational time and stability, given the number and complexity of models, with an exponential covariance function.[26, 27] We use an auto-regressive specification with a single autoregressive parameter *ρ* to allow for temporal correlation.

For the model parameters we specify weakly informative prior distributions, which provide a degree of regularisation and computational stability by limiting the parameters to a plausible range while not being informative within this range.

For the sensitivity and specificity we reviewed previous studies on the performance of RDTs for the different pathogens and specified Beta prior distributions on this basis. The distributions we used are reported in Table 1.

## RESULTS

### Prevalence of enteric pathogens

In total we surveyed and collected stools from 396 infants in round 1 and 181 infants in round 2. Table 2 reports the estimated camp-wide prevalence of the different pathogens. Across both rounds, 62% of infants tested positive for at least one pathogen, with 34% testing positive for two or more. The most common pathogens in both rounds were Campylobacter, Giardia, and Cryptosporidium, all with prevalence over 10% in both rounds of the study. Table 3 reports the proportion of those who tested positive who reported an episode of diarrhoea in the preceding 24 hours. In all cases only a small minority of test positives had had diarrhoea.

**Table 2.** Estimated prevalence of the pathogens from an uncorrected estimator and an estimator corrected for the sensitivity and specificity of the RDTs

| Pathogen | Round |  |  |  |
| --- | --- | --- | --- | --- |
|  | 1 (Jan-Mar 2021) |  | 2 (Sep-Oct 2021) |  |
|  | Uncorrected | Corrected | Uncorrected | Corrected |
| *E. Coli* O157 | 1.7 (0.8, 2.9) | 1.1 (0.1, 2.6) | 2.7 (1.1, 4.9) | 1.9 (0.2, 4.5) |
| *Cryptosporidium* | 14.8 (11.9, 17.8) | 14.1 (9.4, 18.6) | 14.2 (10.2, 18.5) | 13.0 (7.9, 18.4) |
| *Giardia* | 19.4 (16.1, 22.7) | 18.6 (10.8, 26.4) | 26.8 (21.7, 32.2) | 27.4 (18.3, 37.8) |
| *Shigella* | 5.5 (3.8, 7.5) | 5.3 (2.4, 8.3) | 2.2 (0.8, 4.3) | 1.8 (0.2, 4.3) |
| *Campylobacter* | 22.1 (18.7, 25.6) | 22.3 (14.9, 30.4) | 20.7 (15.9, 25.4) | 19.7 (11.2, 28.8) |
| *Rotavirus* | 3.0 (1.8, 4.5) | 2.3 (0.4, 4.2) | 1.6 (0.4, 3.4) | 1.2 (0.1, 3.1) |
| *Norovirus* | 1.3 (0.5, 2.3) | 0.9 (0.1, 2.0) | 0.5 (0.0, 1.6) | 0.5 (0.0, 1.6) |
| *Adenovirus* | 1.2 (0.5, 2.3) | 0.9 (0.1, 2.0) | 0.5 (0.0, 1.7) | 0.6 (0.0, 1.7) |

**Table 3.** Proportion of those who test positive for each pathogen whose stool was diarrhoea

| Pathogen | Round 1 |  | Round 2 |  |
| --- | --- | --- | --- | --- |
|  | Test positive, n | Reporting diarrhoea | Test positive, n | Reporting diarrhoea |
| *E. Coli* O157 | 6 | 0% | 4 | 25% |
| *Cryptosporidium* | 58 | 0% | 25 | 4% |
| *Giardia* | 76 | 0% | 48 | 6% |
| *Shigella* | 21 | 5% | 3 | 0% |
| *Campylobacter* | 87 | 3% | 37 | 8% |
| *Rotavirus* | 11 | 0% | 2 | 0% |
| *Norovirus* | 4 | 25% | 0 | 0% |
| *Adenovirus* | 16 | 6% | 6 | 0% |

### Geospatial mapping

Figures 3 to 5 show the geospatial model outputs for Campylobacter, Giardia, and Adenovirus, respectively (all other outputs are shown in the Supplementary Information). For Campylobacter (Figure 3) there is evidence of raised prevalence in the North-West of the camp where the settlement density and water chlorination is highest, with predicted prevalence 10 percentage points higher than other areas of the camp. The probability that prevalence was above 25% was 60% and 70% in rounds 1 and 2, respectively. For Giardia (Figure 4), there was evidence of clustering of cases in different locations unexplained by observed covariates, particularly in round 2. There was a high probability that the prevalence of Adenovirus (Figure 5) was lower in the chlorinated areas.

**Figure 3.**
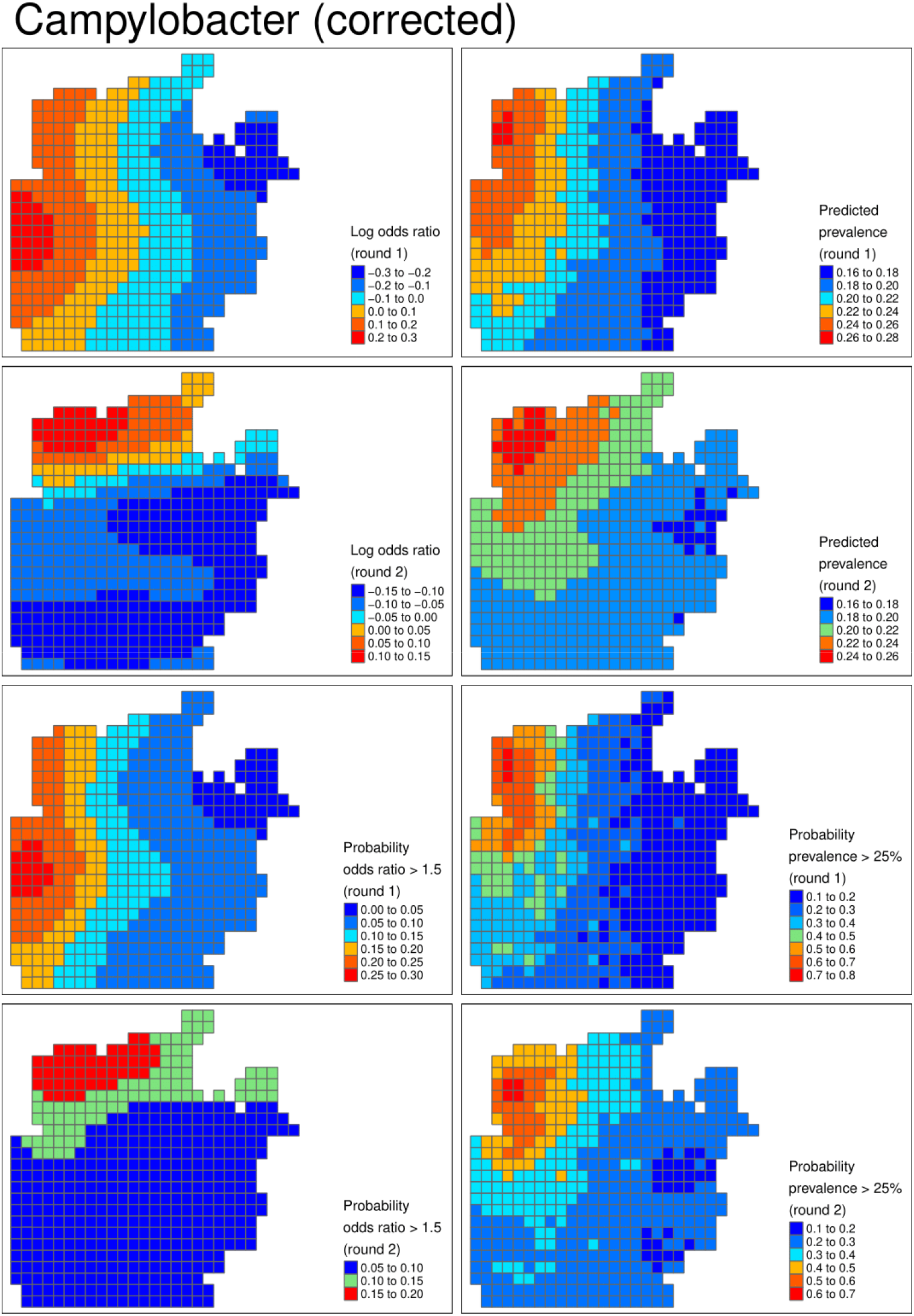
Shigella (corrected). From left to right, from top row to bottom row: log odds ratio describing the latent risk in round 1, predicted prevalence in round 1, log odds ratio in round 2, predicted prevalence in round 2, the probability the odds ratio exceeded 1.5 in round 1, the probability the prevalence exceeded 8% in round 1, and the bottom row is the respective probabilities for round 2.

**Figure 4.**
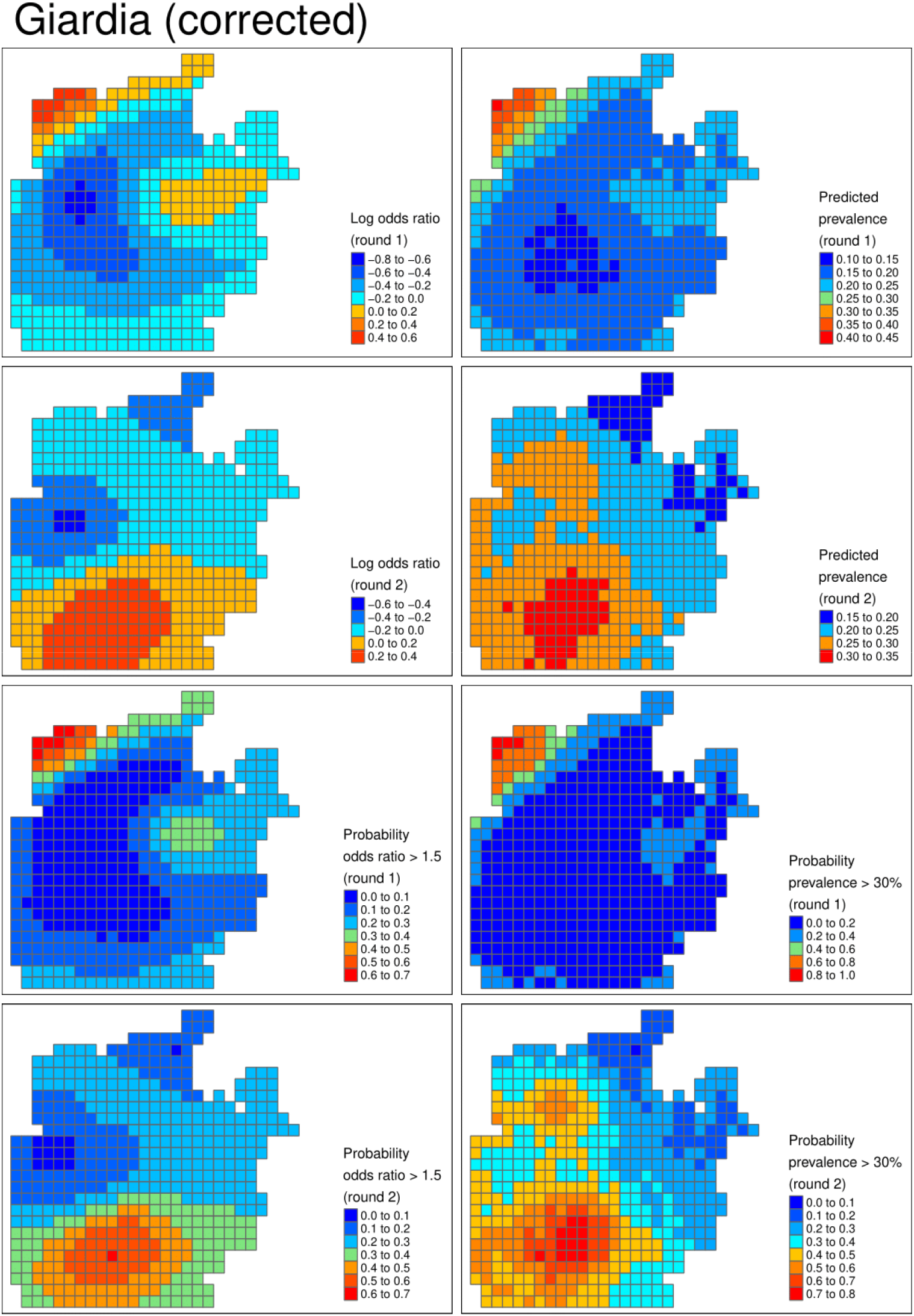
Giardia (corrected). From left to right, from top row to bottom row: log odds ratio describing the latent risk in round 1, predicted prevalence in round 1, log odds ratio in round 2, predicted prevalence in round 2, the probability the odds ratio exceeded 1.5 in round 1, the probability the prevalence exceeded 30% in round 1, and the bottom row is the respective probabilities for round 2.

**Figure 5.**
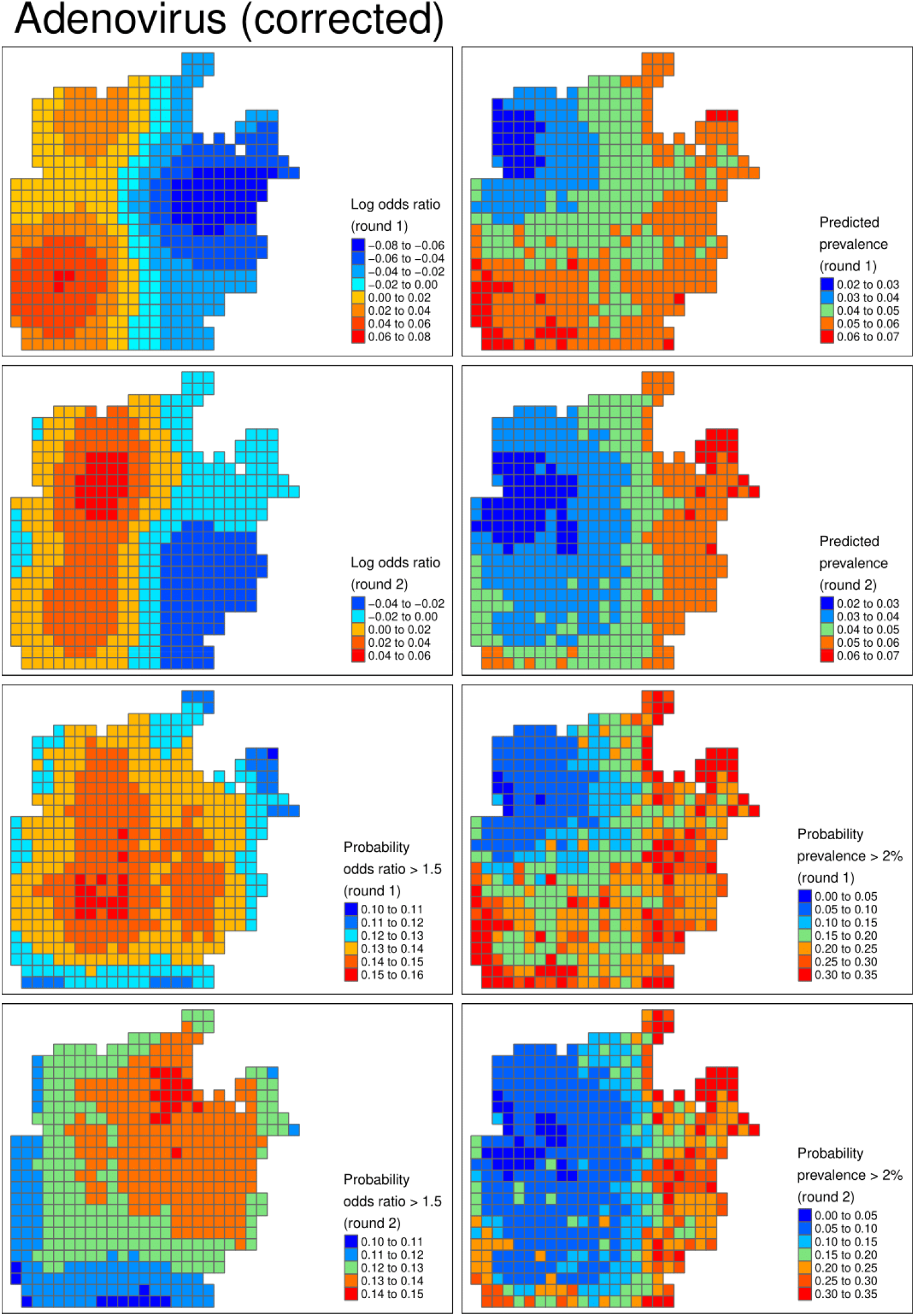
Adenovirus (corrected). From left to right, from top row to bottom row: log odds ratio describing the latent risk in round 1, predicted prevalence in round 1, log odds ratio in round 2, predicted prevalence in round 2, the probability the odds ratio exceeded 1.5 in round 1, the probability the prevalence exceeded 2% in round 1, and the bottom row is the respective probabilities for round 2.

Table 4 reports the model parameters: chlorine was highly predictive of lower prevalence of for all the viruses and Shigella. Comparing all the pathogens tested, all three viruses and Shigella displayed a high probability of reduced prevalence in the area with higher levels of water chlorination. The opposite relationship was predicted for *E. coli*, Campylobacter, and to a lesser extent Cryptosporidium.

**Table 2.** Posterior mean and 95% credible interval of model a parameters from both the uncorrected and corrected models.

| Parameter | Pathogen |  |  |  |  |  |  |  |
| --- | --- | --- | --- | --- | --- | --- | --- | --- |
|  | *E. Coli O157* | *Cryptosporidium* | *Giardia* | *Shigella* | *Campylobacter* | *Rotavirus* | *Norovirus* | *Adenovirus* |
| *Uncorrected* |  |  |  |  |  |  |  |  |
| *Intercept* | -4.62<br>(-6.39, -3.15) | -1.59<br>(-2.28, -0.89) | -1.39<br>(-2.15, -0.56) | -3.78<br>(-5.06, -2.56) | -1.48<br>(-2.18, -0.71) | -4.03<br>(-5.58, -2.61) | -6.80<br>(-9.80, -4.18) | -2.85<br>(-3.92, -1.83) |
| *Population density* | 0.04<br>(-0.45, 0.60) | -0.20<br>(-0.40, 0.00) | -0.04<br>(-0.23, 0.15) | 0.42<br>(0.03, 0.82) | 0.03<br>(-0.16, 0.21) | 0.14<br>(-0.33, 0.65) | 0.69<br>(-0.15, 1.60) | -0.05<br>(-0.37, 0.30) |
| *Chlorine* | 0.23<br>(-0.41, 0.85) | 0.24<br>(-0.06, 0.54) | 0.24<br>(-0.17, 0.66) | -0.61<br>(-1.11, -0.11) | 0.06<br>(-0.29, 0.36) | -0.22<br>(-0.83, 0.37) | -0.46<br>(-1.38, 0.45) | -0.28<br>(-0.75, 0.16) |
| *Length scale* | 1.58<br>(0.12, 3.72) | 1.72<br>(0.17, 3.68) | 0.81<br>(0.07, 2.68) | 1.36<br>(0.08, 3.38) | 1.48<br>(0.16, 3.68) | 1.68<br>(0.20, 3.63) | 1.50<br>(0.10, 3.59) | 1.58<br>(0.09, 3.74) |
| *Autoregressive parameter* | 0.03<br>(-0.87, 0.88) | 0.02<br>(-0.87, 0.86) | 0.10<br>(-0.80, 0.89) | -0.05<br>(-0.88, 0.85) | 0.10<br>(-0.82, 0.90) | -0.01<br>(-0.86, 0.85) | 0.03<br>(-0.85, 0.89) | 0.06<br>(-0.82, 0.89) |
| *Corrected* |  |  |  |  |  |  |  |  |
| *Intercept* | -6.66<br>(-11.40, -3.38) | -1.63<br>(-2.54, -0.77) | -1.49<br>(-2.91, -0.20) | -4.70<br>(-7.77, -2.55) | -1.68<br>(-2.88, -0.40) | -6.17<br>(-10.92, -2.96) | -7.48<br>(-11.94, -3.94) | -5.52<br>(-11.08, -1.94) |
| *Population density* | -0.55<br>(-1.99, 0.83) | -0.23<br>(-0.49, 0.00) | -0.04<br>(-0.34, 0.24) | 0.58<br>(-0.11, 1.41) | 0.07<br>(-0.22, 0.36) | -0.24<br>(-1.61, 0.85) | -0.17<br>(-1.75, 1.16) | -0.42<br>(-1.68, 0.68) |
| *Chlorine* | 0.01<br>(-1.64, 1.61) | 0.27<br>(-0.11, 0.66) | 0.34<br>(-0.21, 0.94) | -0.88<br>(-1.92, -0.05) | 0.09<br>(-0.33, 0.51) | -0.39<br>(-1.79, 1.00) | -0.41<br>(-1.85, 0.97) | -0.47<br>(-1.76, 0.92) |
| *Length scale* | 1.53<br>(0.12, 3.65) | 1.66<br>(0.12, 3.64) | 0.82<br>(0.10, 2.65) | 1.47<br>(0.10, 3.58) | 1.42<br>(0.15, 3.65) | 1.53<br>(0.11, 3.61) | 1.45<br>(0.09, 3.57) | 1.45<br>(0.06, 3.50) |
| *Autoregressive parameter* | 0.00<br>(-0.88, 0.86) | 0.04<br>(-0.83, 0.89) | 0.03<br>(-0.83, 0.87) | -0.04<br>(-0.89, 0.86) | 0.09<br>(-0.85, 0.92) | 0.00<br>(-0.87, 0.89) | -0.01<br>(-0.88, 0.86) | -0.01<br>(-0.85, 0.86) |
| *Sensitivity* | 0.85<br>(0.70, 0.96) | 0.90<br>(0.76, 0.98) | 0.74<br>(0.56, 0.88) | 0.85<br>(0.69, 0.96) | 0.73<br>(0.55, 0.88) | 0.95<br>(0.85, 1.00) | 0.95<br>(0.86, 1.00) | 0.90<br>(0.78, 0.98) |
| *Specificity* | 0.98<br>(0.97, 0.99) | 0.98<br>(0.94, 1.00) | 0.94<br>(0.89, 0.98) | 0.99<br>(0.97, 1.00) | 0.94<br>(0.88, 0.98) | 0.98<br>(0.97, 1.00) | 0.99<br>(0.99, 1.00) | 0.97<br>(0.95, 0.99) |

Model outputs from uncorrected models were qualitatively similar to the corrected model. The uncorrected models suggested higher certainty around the presence of high prevalence areas than the uncorrected models. For example, in round 2 for Giardia, the clustering appears more certain with lower probability of high prevalence in the area around the high prevalence area. Similarly, the credible intervals of the parameters were narrower in the uncorrected models (Table 3).

## DISCUSSION

### Use of RDTs

We used RDTs to survey and map the prevalence of eight enteric pathogens in a FDRPM camp in Bangladesh. The motivation for using RDTs is that they have a lower price, can expedite results, can be used in the field with relatively little training, and obviate the need to transport samples. In previous work in Cox’s Bazar, we collected stool samples in the same manner, but then froze them and shipped them to a laboratory in Dhaka where we then used an appropriate lab-based methods to identify pathogens.[28] We estimated the cost per sample was at least 80% lower with the RDTs than in our earlier study. The field workers in the current study were recruited locally and provided with training and the appropriate materials, which further provided a useful link between the research and the population. Thus, in terms of feasibility, the RDTs were successful.

### Correction and effects

The epistemic cost of using RDTs to collect epidemiological outcomes is the large increase in classification error owing to the imperfect sensitivity and specificity. Our statistical approach incorporated terms for these parameters to allow for the additional uncertainty following previous work in this area.[8, 9] Our results showed that ignoring the error would result in overconfidence and bias in the results, both in terms of the crude prevalence and its spatial and temporal distribution. However, meaningful and interpretable results could be obtained from our corrected models. A key weakness of this study is that we did not have a Gold standard, from which we could estimate the “true” spatial distribution of cases and compare to our results. However, the cost to obtain sufficient samples to estimate these geospatial models was prohibitive. We aimed to use all available evidence on the sensitivity and specificity of the tests to inform our priors for the parameters in the model.

### Prevalence of pathogens and distribution

Our overall prevalence estimates were highly comparable to our previous study in this camp where PCR-based methods were used.[28] Our results suggest that recent efforts in the camp to chlorinate the water were successful in reducing the prevalence of particularly viruses. However, *E*.*coli*, Campylobacter, and Cryptosporidium were predicted to have higher prevalence in the high density region of the camp. Two mechanisms might explain these observations. First, transmission may be linked to food or environmental contamination, facilitated by higher population densities, rather than water. Second, they may be resistant to lower levels of chlorination. Cryptosporidium is known to be highly chlorine tolerant.[29, 30] One recent study suggested that Campylobacter isolates can survive or be revived from exposure to relatively high concentrations of chlorine,[31] and similar observations have been made for various strains of *E*.*coli*.[32, 33] Our highest recorded value for chlorine was 3 ppm.

### Limitations

There were other limitations to our study. Our fieldwork was significantly affected by Covid-19. We aimed to conduct a survey at the heights of both the wet and dry seasons, as the prevalence of viruses and bacteria vary.[28] However, this was not possible and the two rounds of our survey took place in relatively similar climates. We also aimed to follow up with the same households in rounds 1 and 2 to consider intra-person comparisons and facilitate sampling in round 2. However, many of the infants could not be re-located in round 2 due to significant movement of people within and between camps due to on-going programs to relocate the FDRPMs.

### Diarrhoeal disease

Our results also suggest that the carriage rate of all the organisms was much higher than the rate of symptomatic diarrhoeal illness. The vast majority of test positive stools were not liquid or watery. Many studies evaluating WASH interventions have used self-reported diarrhoea as an outcome. In previous work, we have shown how diarrhoea is poorly associated with enteric infection, irrespective of how diarrhoea is measured.[28] We argue that diarrhoea itself can be seen as an imperfect test of enteric infection, albeit with much poorer diagnostic performance than the RDTs we used in this study. The diagnostic error associated with diarrhoea is unknown and is likely to vary from place to place, meaning an appropriate correction cannot be made, leading to bias in estimators of prevalence and hence the effectiveness of interventions using these measures. One of the arguments for using self-reported diarrhoea is that it is cheap to collect, meaning large samples can be obtained, and it does not require access to expensive specialist labs, equipment, and personnel, which are often not available in resource-poor contexts. In this study, we have demonstrated that RDTs can provide a useful middle-ground, facilitating the collection of data on the prevalence of enteric pathogens while being low cost and possible to use in difficult or isolated environments.

## Data Availability

Given that the data contain precise location information on study participants, we have not made these data freely available, but we will provide the data and code to reproduce the analyses upon reasonable request.

## ETHICS APPROVALS

Ethics approvals were granted by the Biomedical Science Research Ethics Committee at the University of Warwick, Coventry, UK and the Research Ethics Committee at the International Centre for Diarrhoeal Disease Research, Bangladesh, Dhaka, Bangladesh.

## AUTHOR CONTRIBUTIONS

SIW conceived the idea for the study with support from MSI and RTTR. SIW, MSI, RTTR, and RJL designed the study, which was led in Cox’s Bazar by MSI, AKB, MAA, ASGF and BA. SIW conducted the data analysis and produced the first draft of the manuscript. All authors reviewed, revised, and approved subsequent versions of the manuscript.

## SUPPLEMENTARY DATA

Supplementary data are available at *IJE* online.

