## Supplemental Information for "Low cost and real-time surveillance of enteric infection and diarrhoeal disease using rapid diagnostic tests: A pilot study"

**SUPPLEMENTARY INFORMATION**

**Statistical model**

Our statistical model is a geospatial Binomial model whose realisation is observed on the Cartesian area of interest $A\subset\mathbb{R}^{2}$ and at time period $t\in[1,2]$. The data result from a set of dichotomous tests for a given pathogen, the result of a test at location $s\in A$ and time period $t$ is $y\left( s,t \right)\in[0,1]$, and is distributed as

$$y\left( s,t \right)\sim Bernoulli\left( p\left( s,t \right) \right)$$

Where

$$h\left( p\left( s,t \right) \right)=\mu+x\left( s,t \right)\beta+Z\left( s,t \right)$$

where $x\left( s,t \right)$ is a length $q$ vector of spatially and temporally referenced covariates, $\mu$ is an intercept term, $\left\{ Z\left( s,t \right):s\in A,t\in\left[ 1,2 \right] \right\}$ is a latent field, and $h$is the logit function. We use an auto-regressive specification for the latent field:

$$Z\left( s,1 \right)=\Omega_{1}\left( s \right)$$

$$Z\left( s,2 \right)=\rho\Omega_{1}\left( s \right)+\left( \sqrt{1+\rho^{2}} \right)\Omega_{2}\left( s \right)$$

We take a Bayesian approach and specify a Gaussian process as a prior distribution for each $\left\{ \Omega_{t}\left( s \right):s\in A \right\}$, which represents “spatial innovation” and where $\rho$ is the autoregressive parameter. We use a minimally parameterised covariance function for the Gaussian process:

$$cov\left( \Omega_{t}\left( s \right),\Omega_{t}\left( s^{'} \right) \right)=\sigma^{2}f\left( \left| \left| s-s^{'} \right| \right|^{2};\phi\right)$$

where $||.|\left. \right|^{2}$ is the L2-norm. Here we use the squared exponential covariance function:

$$f\left( \left| \left| s-s^{'} \right| \right|^{2};\phi\right)=\exp\left( \frac{-\left( \left| \left| s-s^{'} \right| \right| \right)^{2}}{2\phi} \right)$$

where $\phi$ is the spatial range (or length scale) parameter.

The model as described above is our “uncorrected model” that does not allow for diagnostic error. To allow for the diagnostic test to have imperfect sensitivity and specificity we also specify a second model in which

$$p\left( s,t \right)=\left( 1-Spec \right)+\left( Sens+Spec-1 \right)*h^{-1}\left( \mu\left( s,t \right) \right)$$

where $Spec$ and $Sens$ are the specificity and sensitivity, respectively.

***Prior distributions***

For the model parameters we specify weakly informative prior distributions, which provide a degree of regularisation and computational stability by limiting the parameters to a plausible range while not being informative within this range. In particular, we use standard normal priors for the parameters in the linear predictor except for $\mu\sim N\left( {0,5}^{2} \right)$. We set $\sigma^{2}\sim N\left( 0,1 \right)[0,\infty)$.

For the sensitivity and specificity we reviewed previous studies on the performance of RDTs for the different pathogens and specified Beta prior distributions on this basis. The distributions we used are reported in Table 1.

***Approximation***

Gaussian processes are flexible models for multi-dimensional non-linear functions. However, they are computationally expensive and difficult to estimate, especially with the more complex corrected model in this article. We use the fast and accurate approximation for fully Bayesian Gaussian Processes proposed by Solin and Särkkä.[26, 27] The approximation is based on Laplacian eigenfunctions for stationary covariance function.

***Software***

We used Stan 2.24 for all the models reported in this paper.

**Additional results**

We present below the outputs from the uncorrected and corrected models for all pathogens, the corrected models for Campylobacter, Giardia, and Adenovirus are shown in the main body of the paper.

For each image below the plots show, from left to right, from top row to bottom row: log odds ratio describing the latent risk in round 1, predicted prevalence in round 1, log odds ratio in round 2, predicted prevalence in round 2, the probability the odds ratio exceeded 1.5 in round 1, the probability the prevalence exceeded the threshold value in round 1, and the bottom row is the respective probabilities for round 2.

***Corrected models***

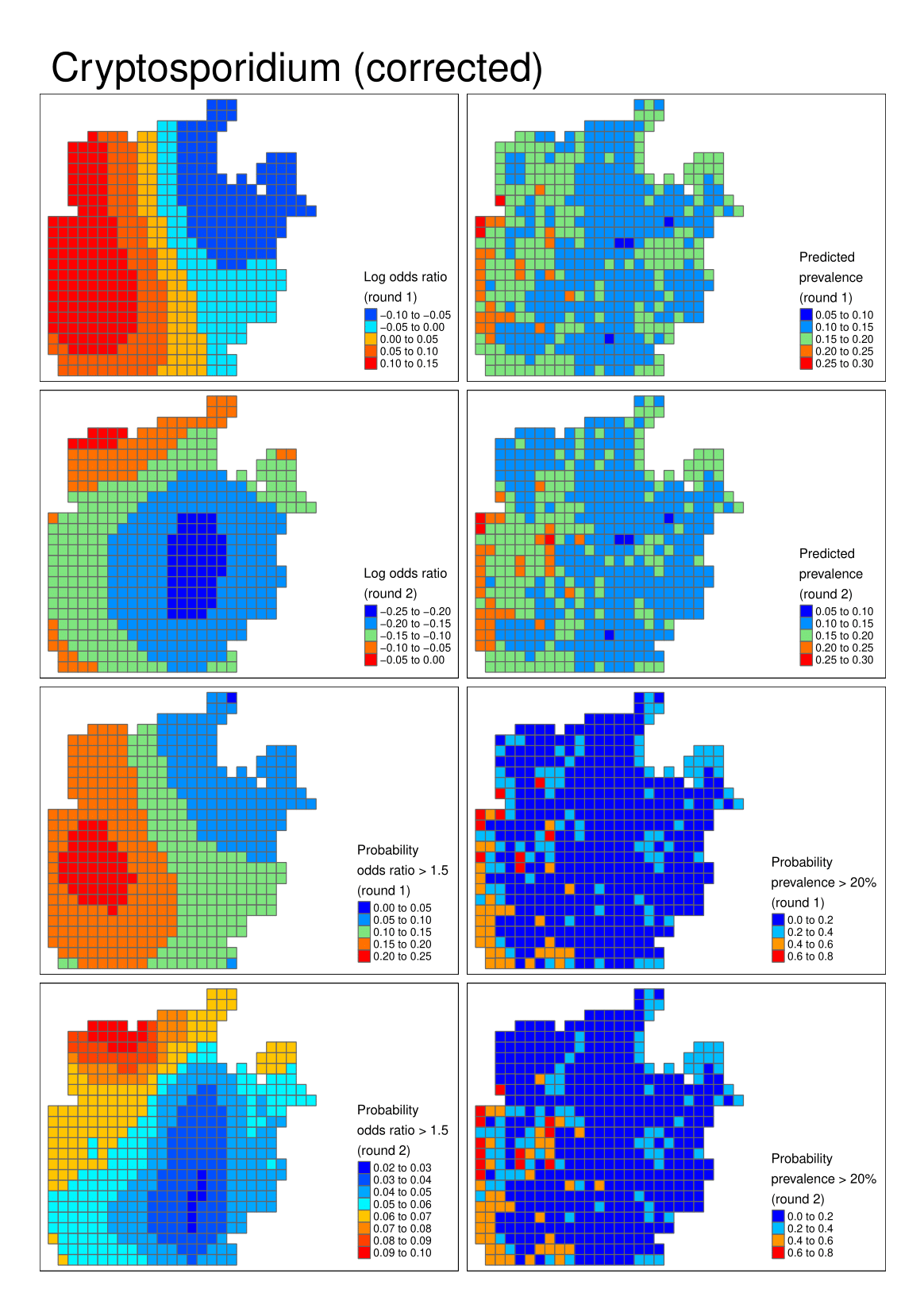

**Figure S1** Cryptosporidium

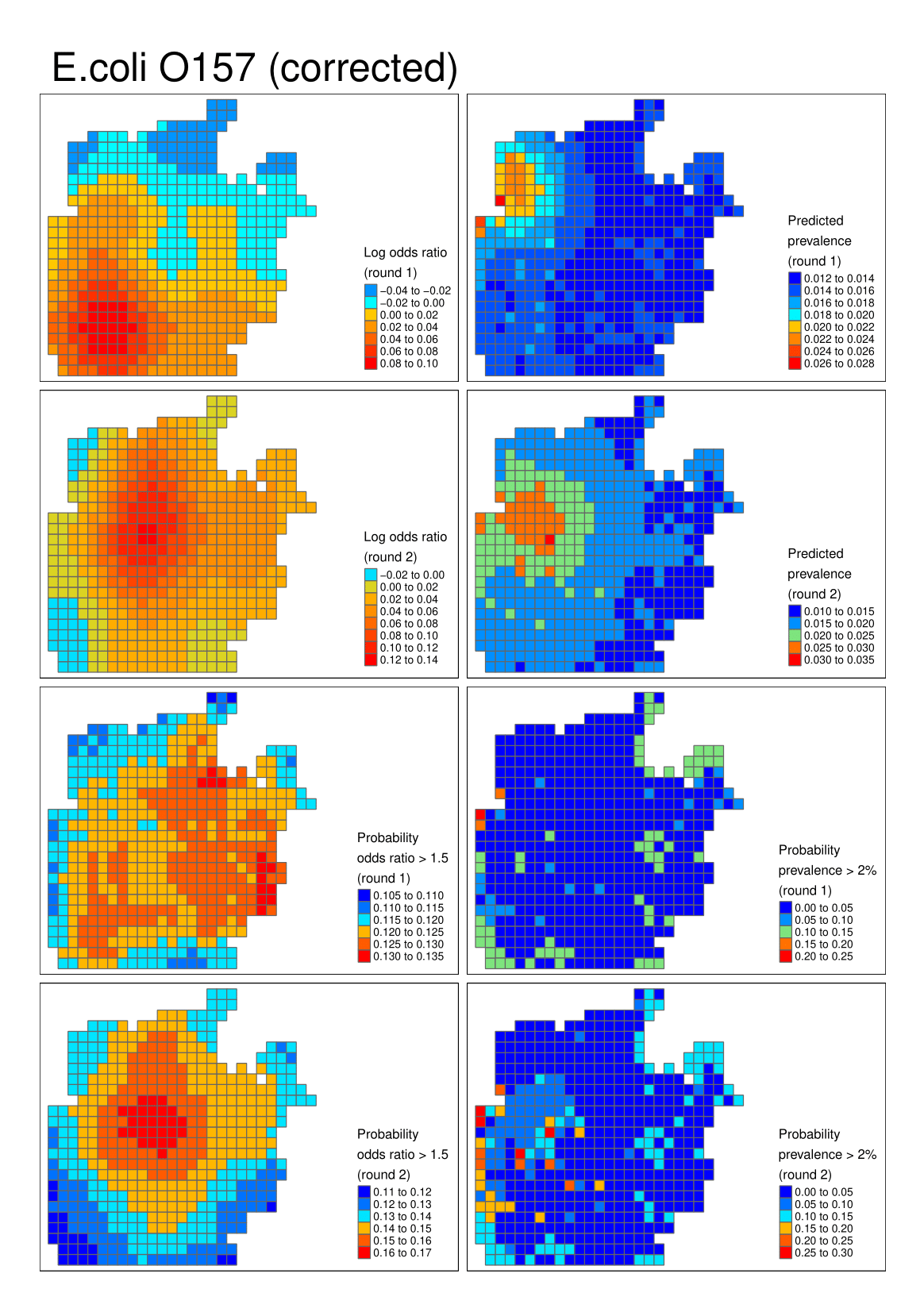

**Figure S2** E.coli O157

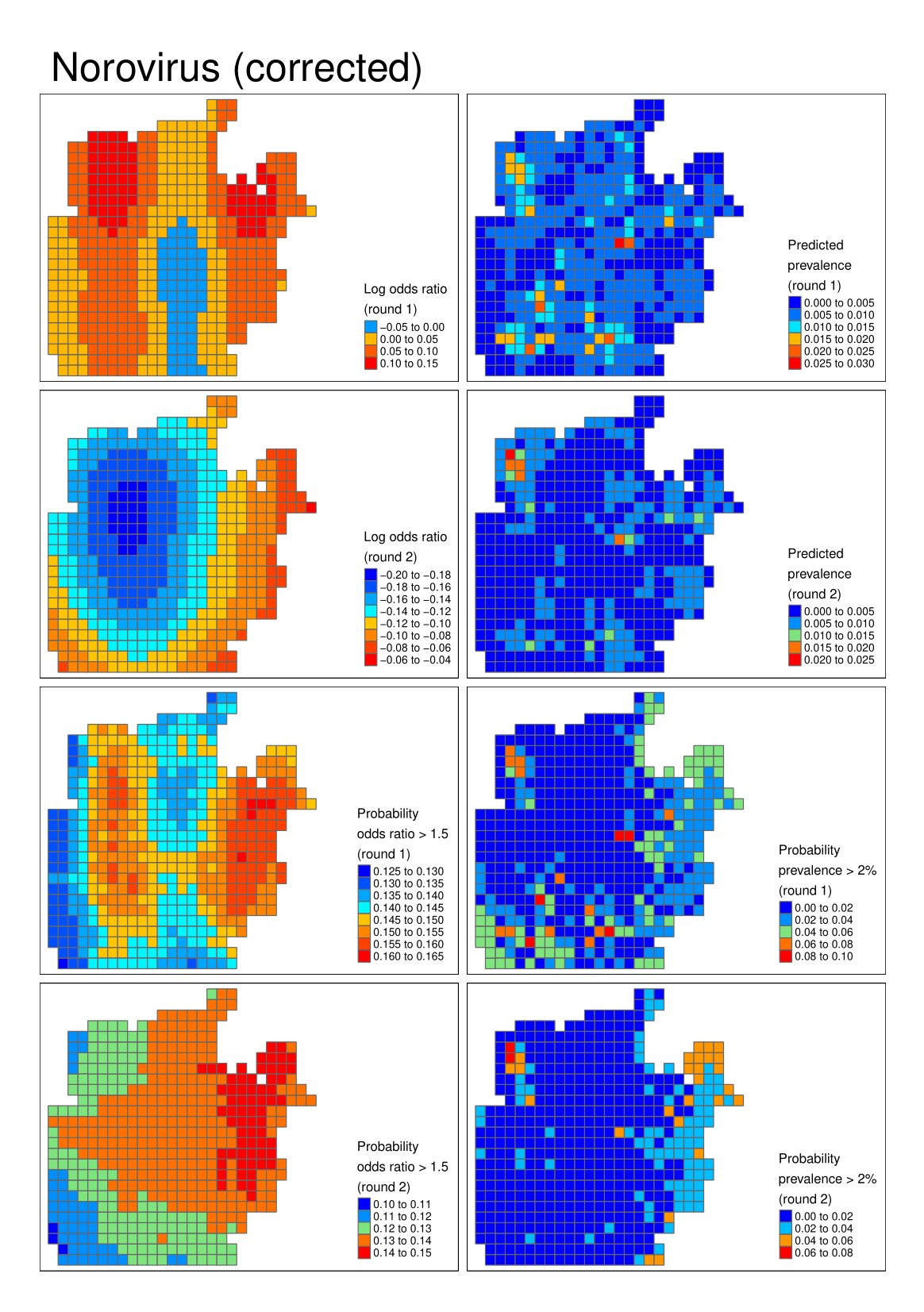

**Figure S3** Norovirus

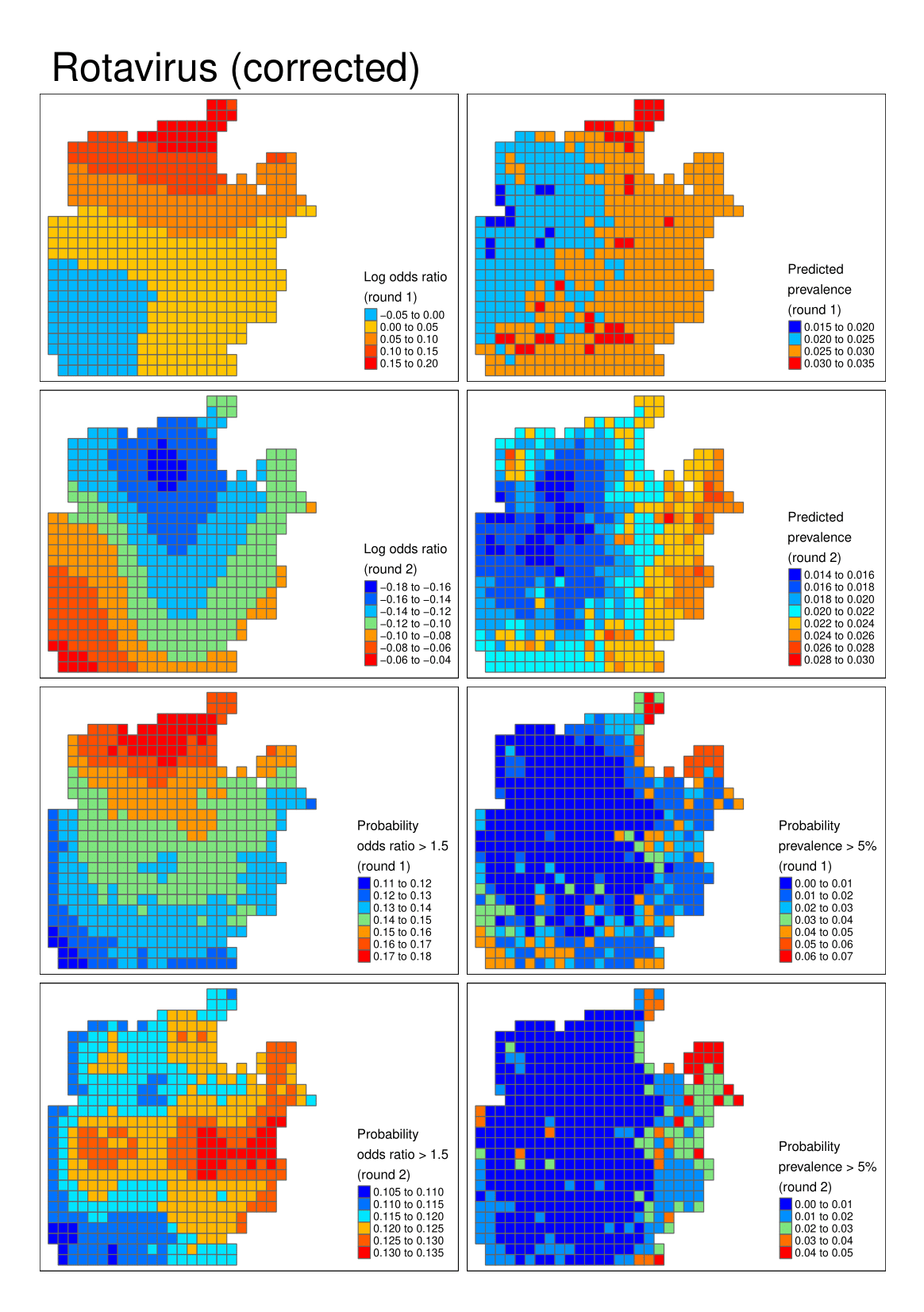

**Figure S4** Rotavirus

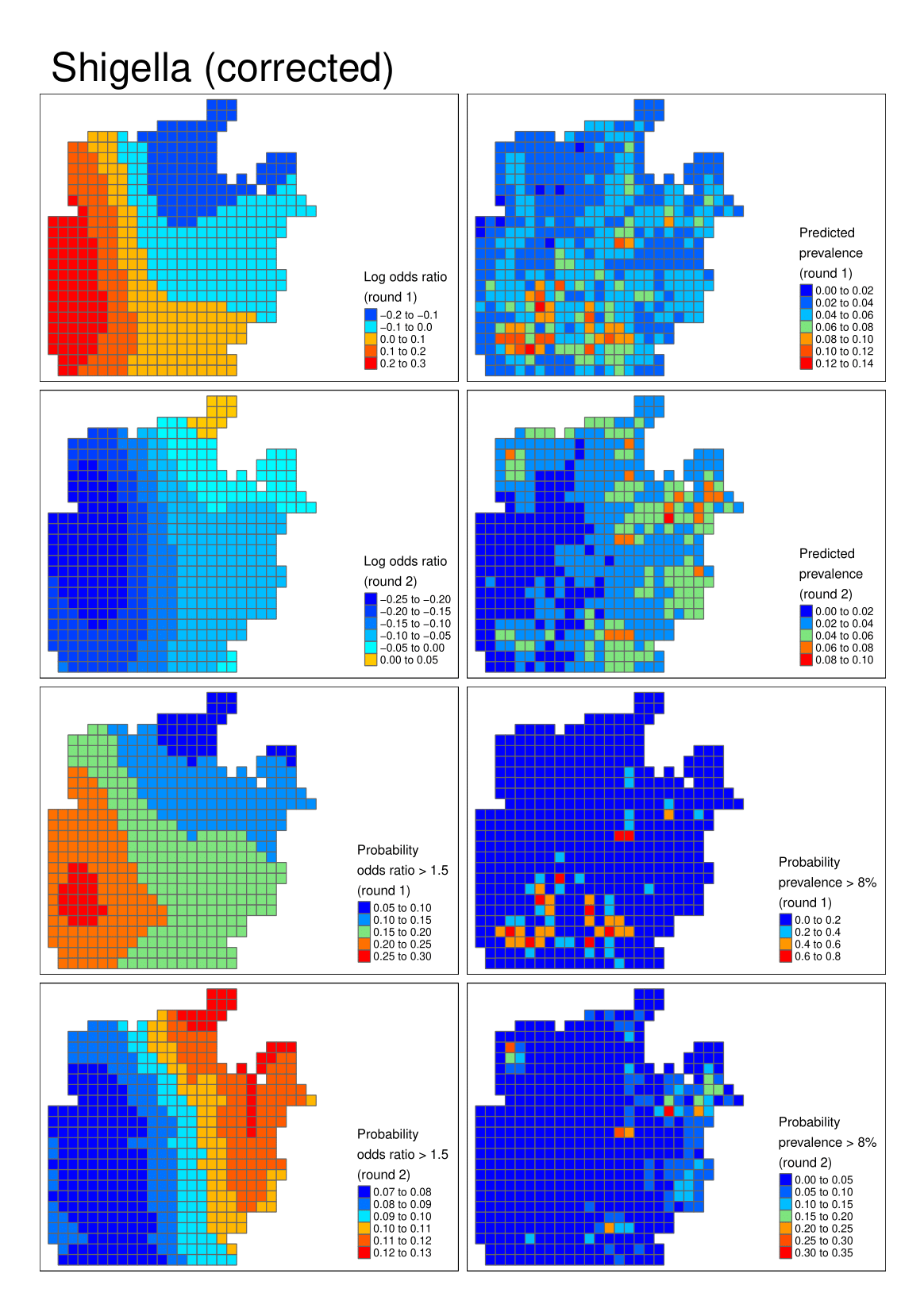

**Figure S5** Shigella (STEC)

***Uncorrected models***

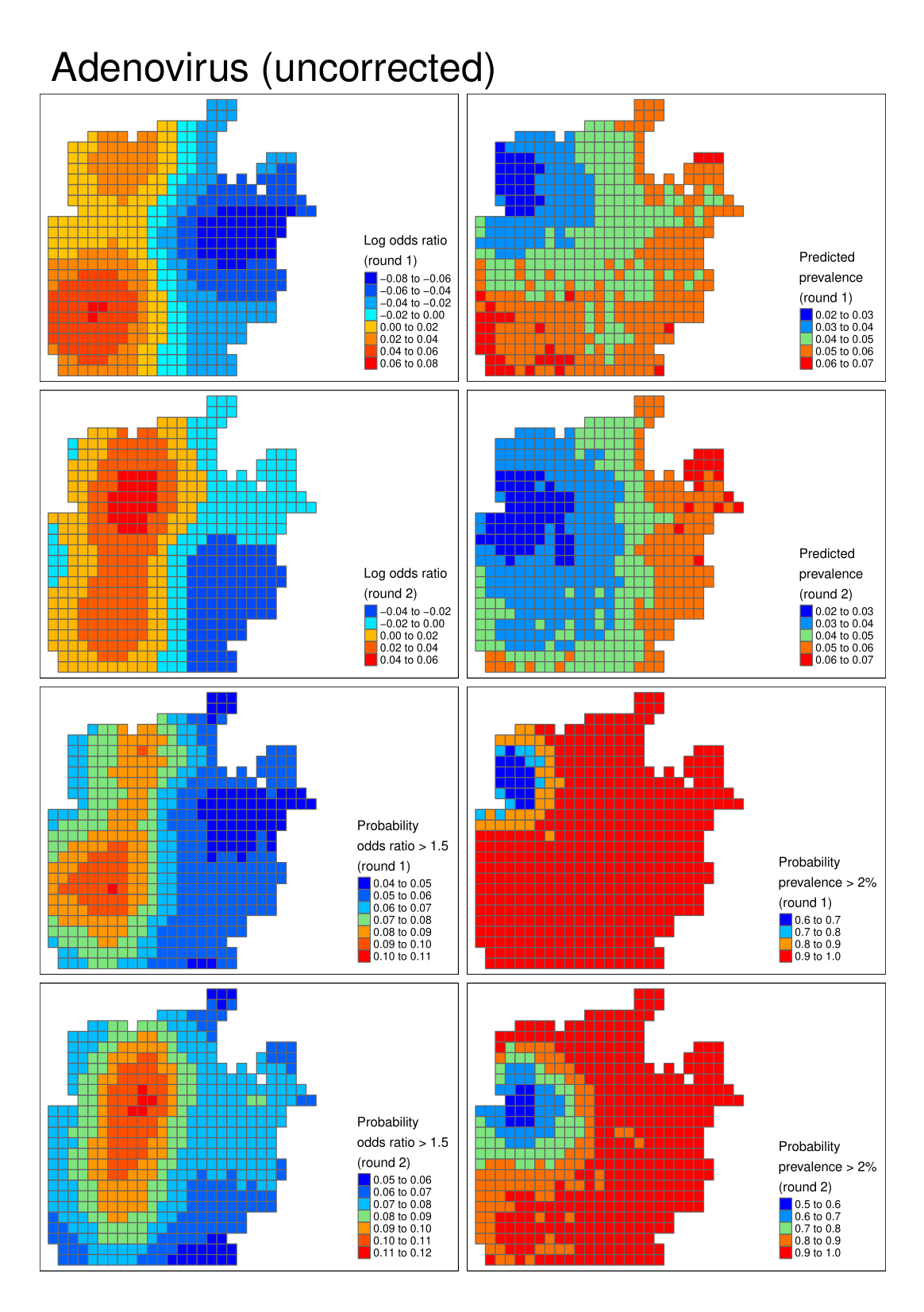

**Figure S6** Adenovirus

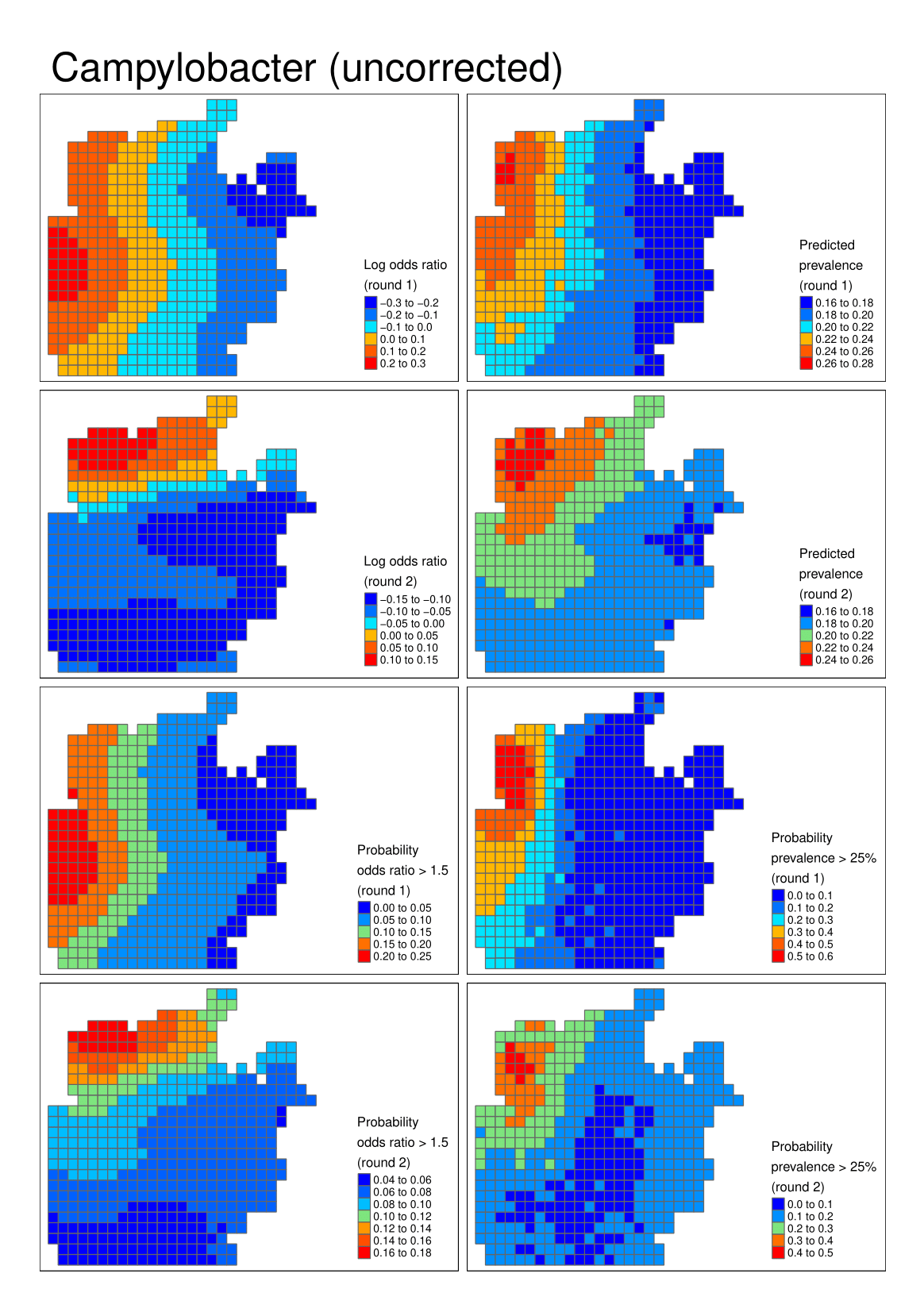

**Figure S7** Campylobacter

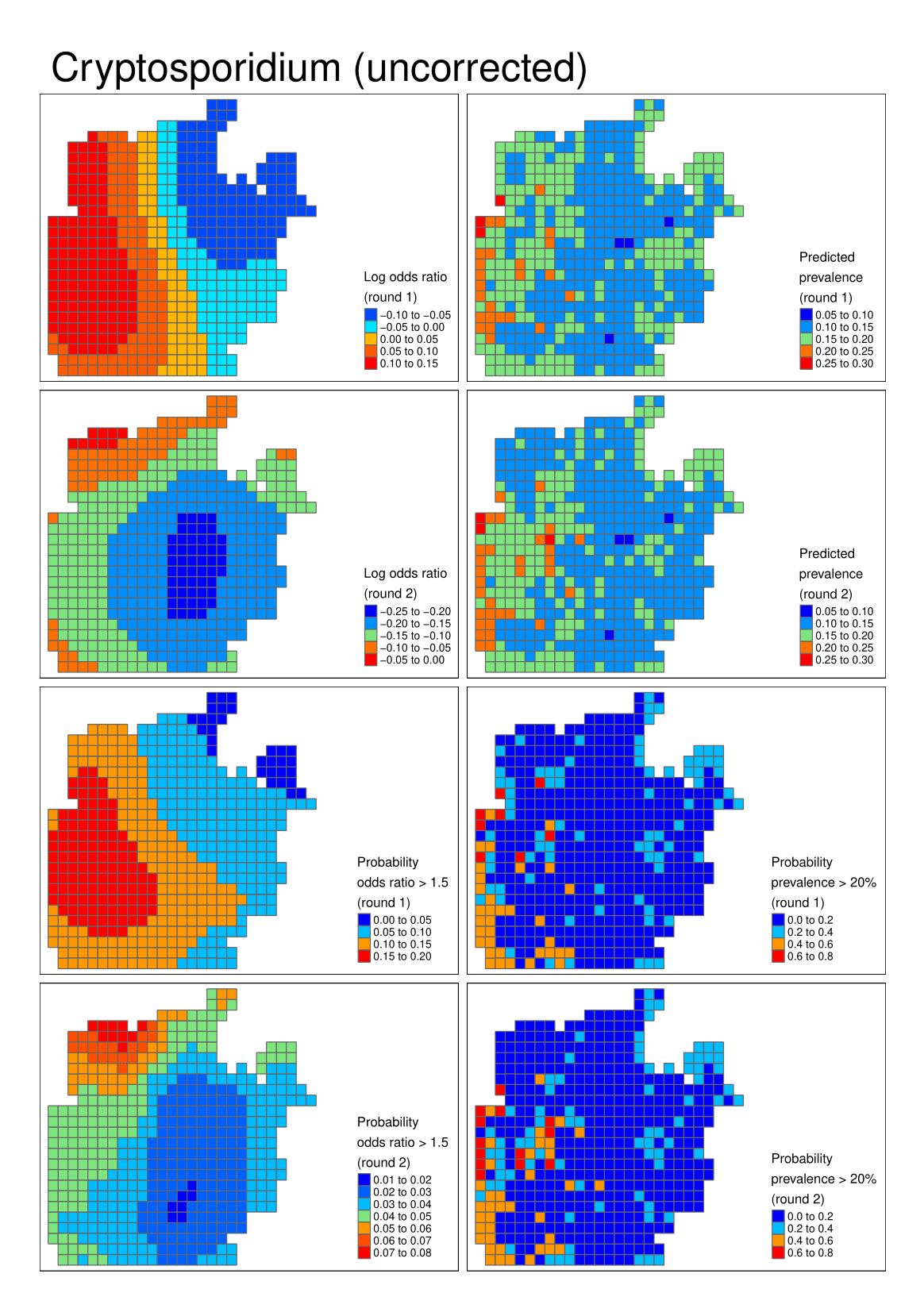

**Figure S8** Cryptosporidium

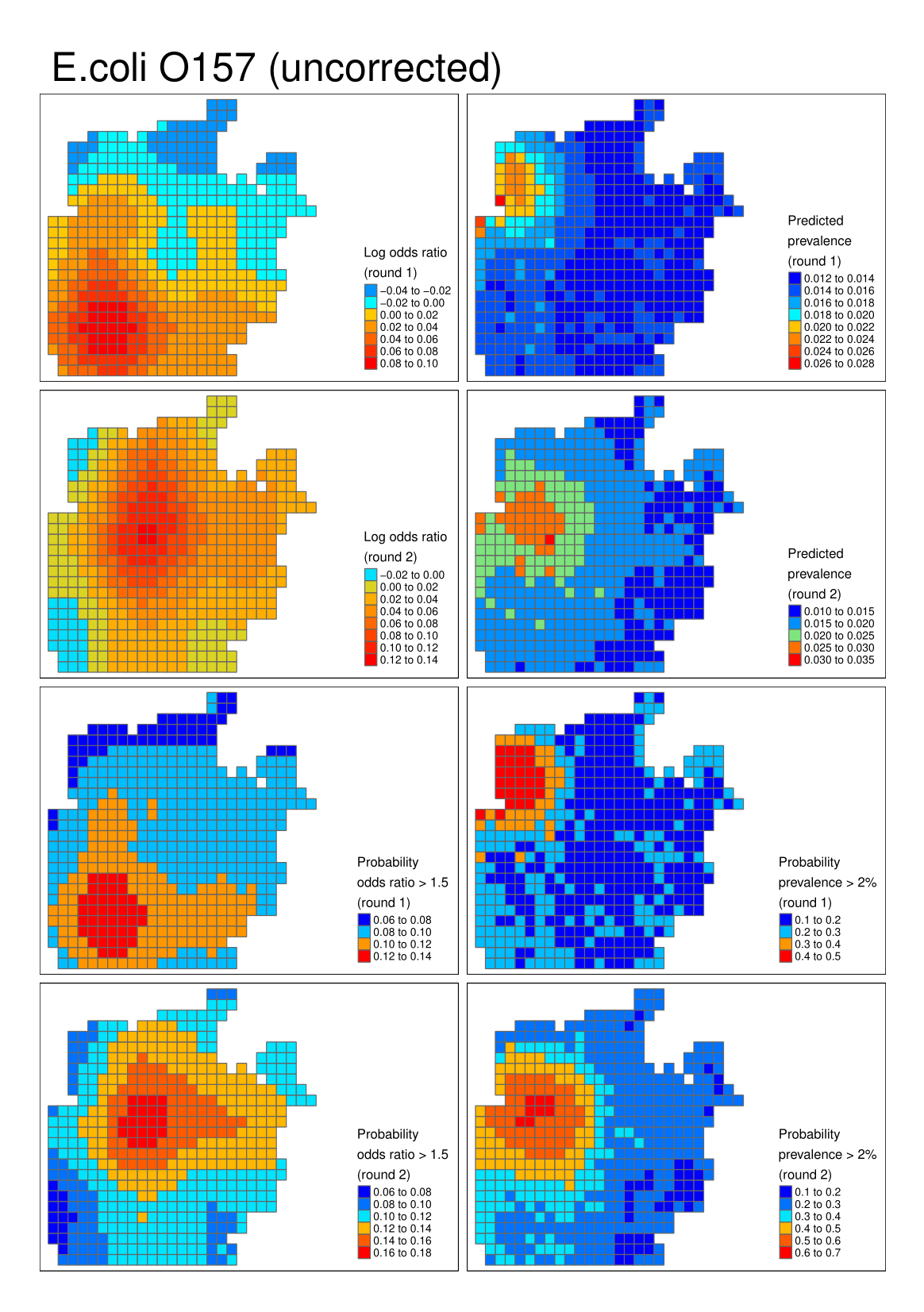

**Figure S9** E.coli O157

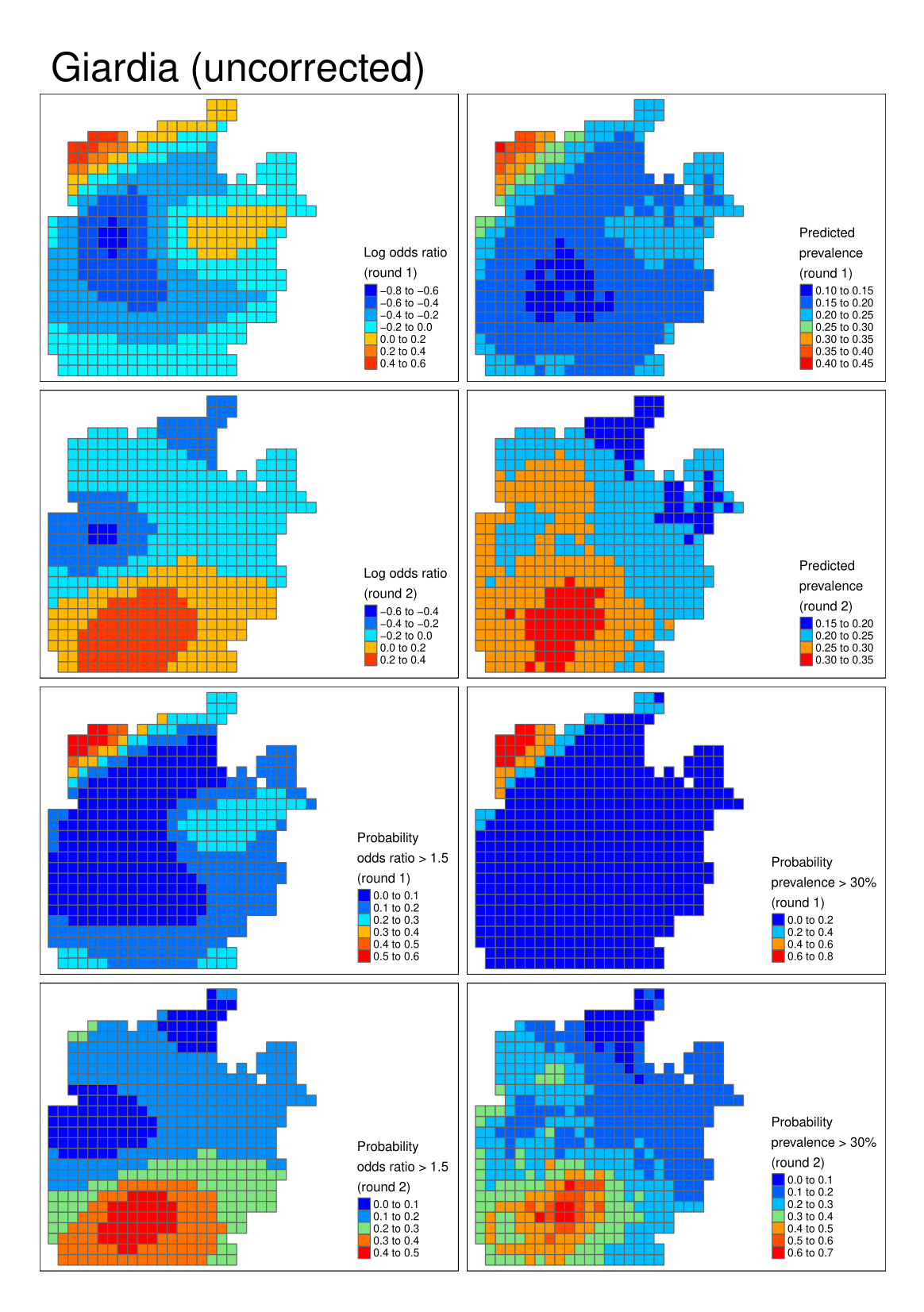

**Figure S10** Giardia

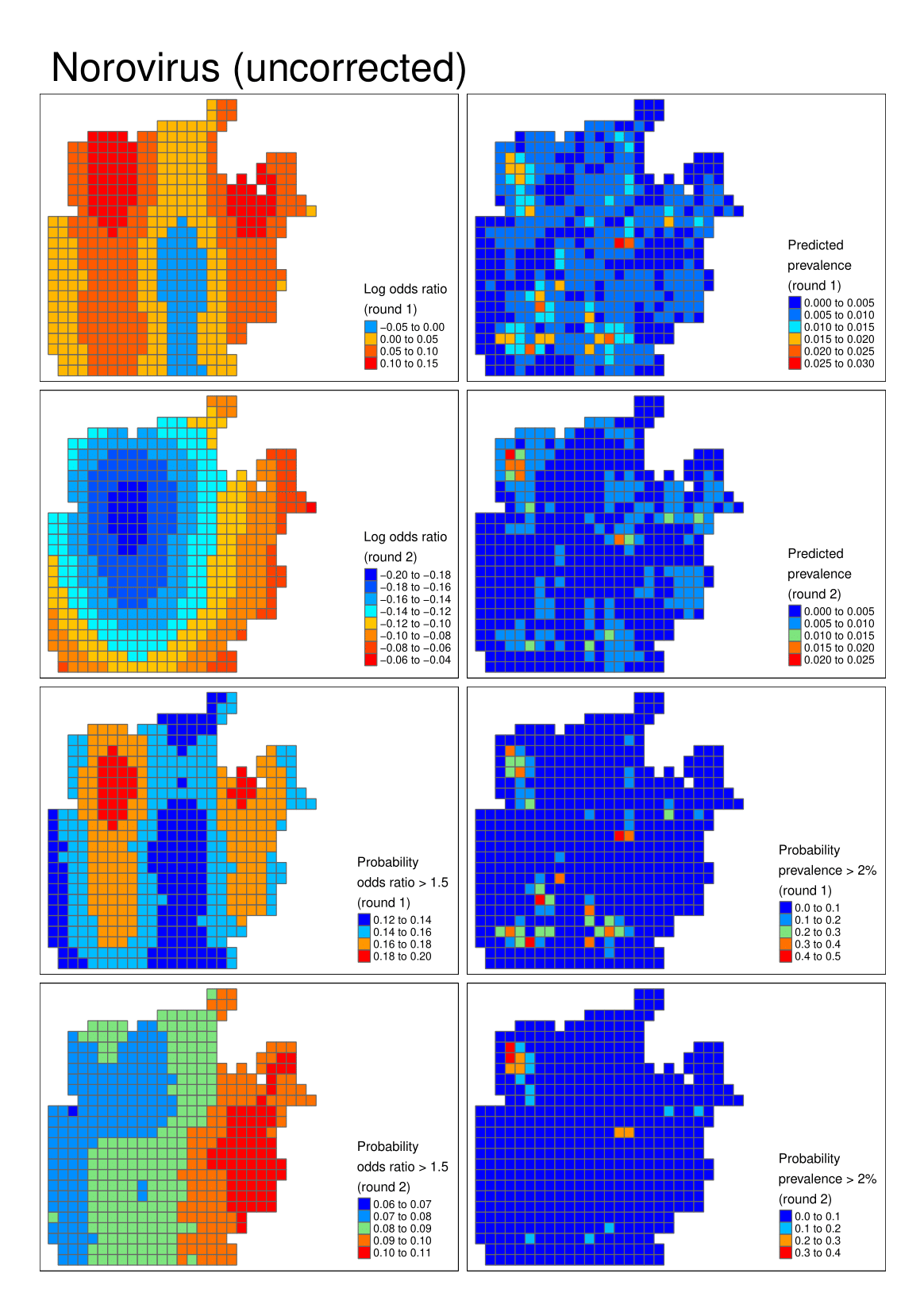

**Figure S11** Norovirus

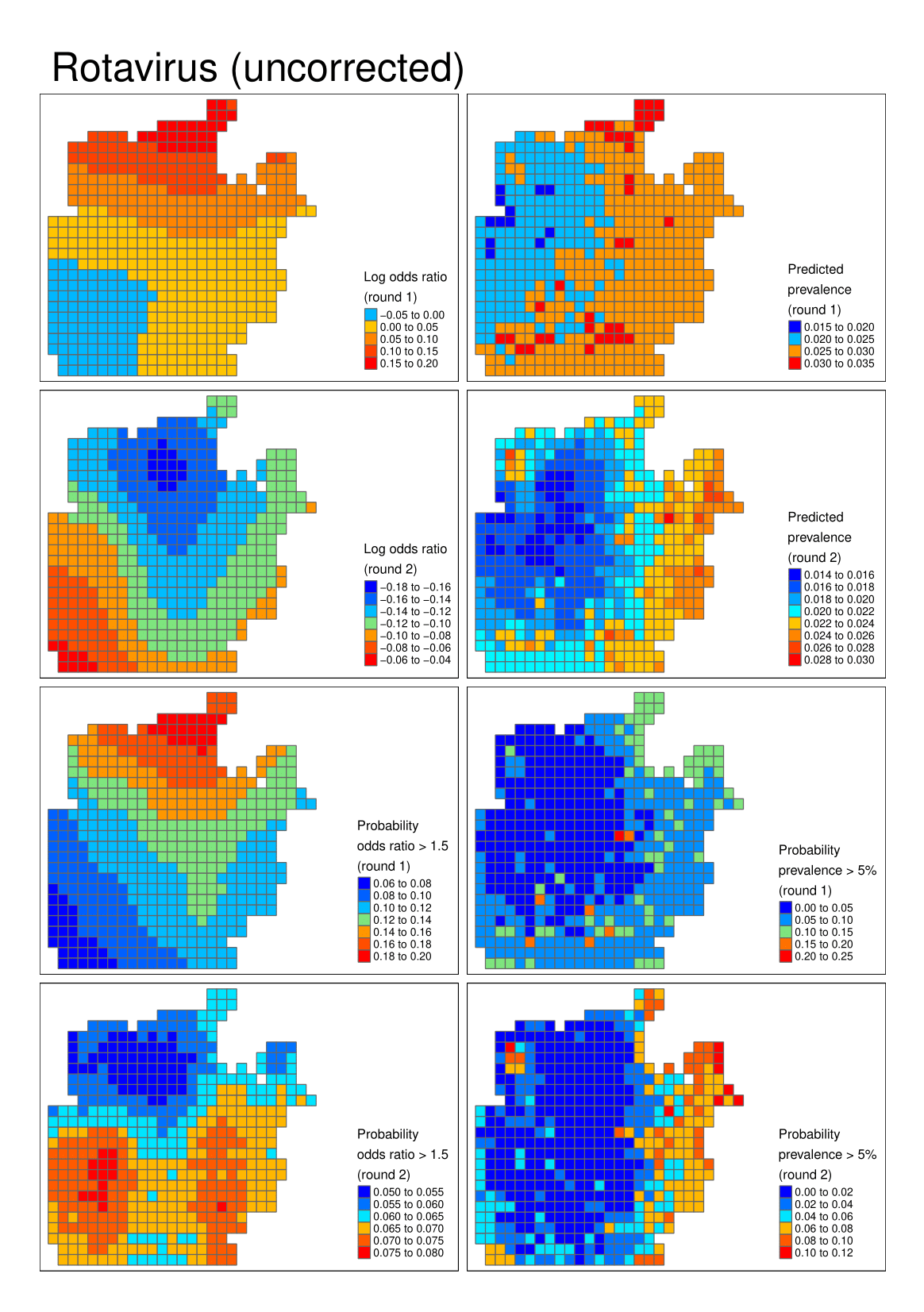

**Figure S12** Rotavirus

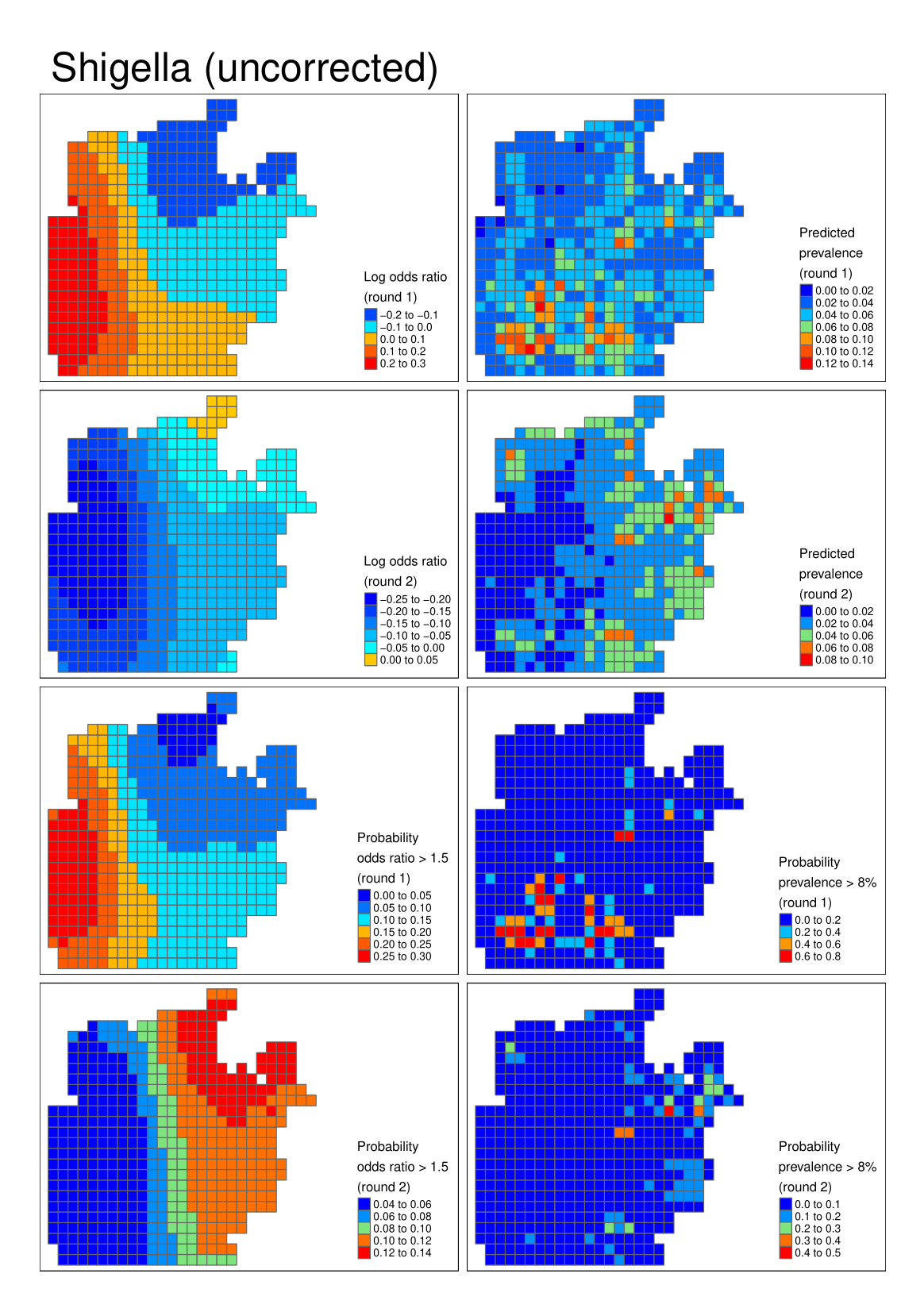

**Figure S13** Shigella (STEC)
